# Detecting salivary host-microbiome RNA signature for aiding diagnosis of oral and throat cancer

**DOI:** 10.1101/2022.07.30.22278239

**Authors:** Guruduth Banavar, Oyetunji Ogundijo, Cristina Julian, Ryan Toma, Francine Camacho, Pedro J. Torres, Lan Hu, Liz Kenny, Sarju Vasani, Martin Batstone, Nevenka Dimitrova, Momchilo Vuyisich, Salomon Amar, Chamindie Punyadeera

## Abstract

**Objectives:** Oral squamous cell carcinoma (OSCC) and oropharyngeal squamous cell carcinoma (OPSCC) are the two major subtypes of head and neck cancer (HNC) that can go undetected resulting in late detection and poor outcomes. We describe the development and validation of a convenient and easy-to-use test, called CancerDetect for Oral & Throat cancer™ (CDOT), to detect markers of OSCC and/or OPSCC within a high-risk population using salivary metatranscriptomics.

**Materials and Methods:** We collected saliva samples from 1,175 unique individuals who were 50 years or older, or adults who had a history of tobacco use. All saliva samples were processed through a metatranscriptomic method to isolate microbial organisms and functions, as well as human transcripts. Of the 1175 samples, 945 were used to train a classifier using machine learning methods, resulting in a salivary RNA metatranscriptomic signature. The classifier was then independently validated on the 230 remaining samples unseen by the classifier, consisting of 20 OSCC (all stages), 76 OPSCC (all stages), and 134 negatives (including 14 pre-malignant).

**Results:** On the validation cohort, the specificity of the CDOT test was 94%, sensitivity was 90% for participants with a histopathological diagnosis of OSCC, and 84.2% for participants with a diagnosis of OPSCC. Similar classification results were observed among people in early stage (stages I & II) vs late stage (stages III & IV) of OSCC and OPSCC.

**Conclusions:** CDOT is a non-invasive test that can be easily administered in dentist offices, primary care centers and specialized cancer clinics for early detection of OPSCC and OSCC. This test, having received breakthrough designation by the US Food and Drug Administration (FDA), will broadly enable early diagnosis of OSCC and OPSCC, saving lives and significantly reducing healthcare expenditure.

## Introduction

Oral cancer is the seventh-most common neoplasm and the ninth most common cause of cancer related death globally [1]. The American Cancer Society estimates about 54,000 new cases of oral cancer, leading to 11,230 deaths, in the United States in 2022 [2]. More than half of oral cancers in the world occur in Asia, in South/Southeast Asia, oral cancer is one of the top three cancers [3]. Oral squamous cell carcinoma (OSCC) is the most common oral cancer, accounting for 2% of all cancers and with a high recurrence rate even with treatment [4]. Oropharyngeal squamous cell carcinoma (OPSCC), commonly known as throat cancer, is currently emerging in the developed world as the leading cause of human papillomavirus (HPV)-driven cancer in the USA, and Australia. While it shares similar etiologic factors with OSCC like smoking history or alcohol consumption, OPSCC is also highly associated with HPV, which makes this cancer biologically and clinically different [5–7].

Survival rates of OPSCC and OSCC patients vary based on stage at the time of diagnosis and disease progression [8]. The five-year overall survival rate in the U.S. for OSCC is 84%, if diagnosed in the early stages of the disease (i.e., Stage I or II). However, more than 70% of OSCC diagnoses are not made until the disease is in stage III or IV. At these later stages, the five-year survival rate, for OSCC specifically, drops to less than 50% [9]. Research has shown that the reasons for late diagnosis are layered and complicated, including under-utilization of dental and primary care, and the lack and poor quality of oral cancer screening in patients that do seek general care [10]. In addition, most importantly, in the earliest, most treatable stages, many oral cancers have little to no symptoms and may not be easily visible [11,12].

The current standard of care for oral cancer screening and diagnosis relies on a physical exam by a healthcare provider, identification of lesion(s), followed by imaging, invasive biopsy and histopathological evaluation. Biopsies will only sample a limited amount of cancer tissue and heterogeneity within the cancer is not accounted for. There are no oral cancer screening guidelines published either from the American Cancer Society, the National Comprehensive Cancer Network (NCCN), or the National Cancer Institute. The only recommendation that exists for oral cancer is in the form of a resolution passed by the American Dental Association in 2019 recommending dentists to conduct routine visual and tactile examinations for oral and oropharyngeal cancer for all patients [13]. Thus, clinicians are left to determine the best practice on their own with no objective criteria or tools for assessing patients. Any abnormalities identified on visual or tactile examination are referred for biopsy. However, only 29.4% of adults in the United States reported ever having received a visual and tactile examination for OSCC or OPSCC [14]. In addition, those patients referred for biopsies go through an invasive and risky procedure with uncertain outcomes based on the specific tissue that was extracted. The most common risk associated with the procedure is a hematoma, or a pocket of blood, which can form and collect at the site of the biopsy. Studies have documented the dissemination of cancer cells into the circulation resulting in an increased risk of metastasis after the incisional biopsy [15]. Moreover, the standard biopsy techniques may not be appropriate for all patients, including those with conditions that preclude the safe use of local anesthetic and those with severe bleeding diathesis or coagulopathies [14].

Even though tobacco consumption, alcohol abuse and poor oral hygiene remain the major risk factors for oral cancer, there has been increasing evidence to suggest that people who are not exposed to these risk factors are also affected. Dysbiosis in the oral microbiome leads to a chronic inflammatory state, suppresses anti-tumor immunity, and leads to the creation of novel mutagens [16]. One of the examples supporting this evidence is periodontitis, which is associated with an increased risk for cancer and poor survival in many studies [17,18]. Streptococcus, Fusobacterium, Capnocytophaga, Prevotella, among other bacteria are shown to be increased in OSCC [19–22]. Changes in microbiota have been observed in throat cancer patients as well [16]. This provides the scientific evidence to further explore microbial organisms and functions in the saliva as a means of developing a tool to evaluate oral and throat cancers.

Previously we developed a classifier for the detection of OSCC using only microbial expression on a smaller cohort [23]. In this current study, we incorporate both OSCC and OPSCC, include human gene expression in addition to microbial expression, and expand the studied cohort significantly. The resulting test, CancerDetect for Oral & Throat Cancer™ [CDOT], built using salivary metatranscriptomics and validated with an independent cohort, was granted breakthrough designation by the Food and Drug Administration (FDA) in April 2021.

## Test Description

We have developed a simple cancer detection test as shown in Figure 1, consisting of the following elements: (i) Sample collection / transport, (ii) Lab Processing (iii) Data Processing, and Test Report.

**Figure 1:**
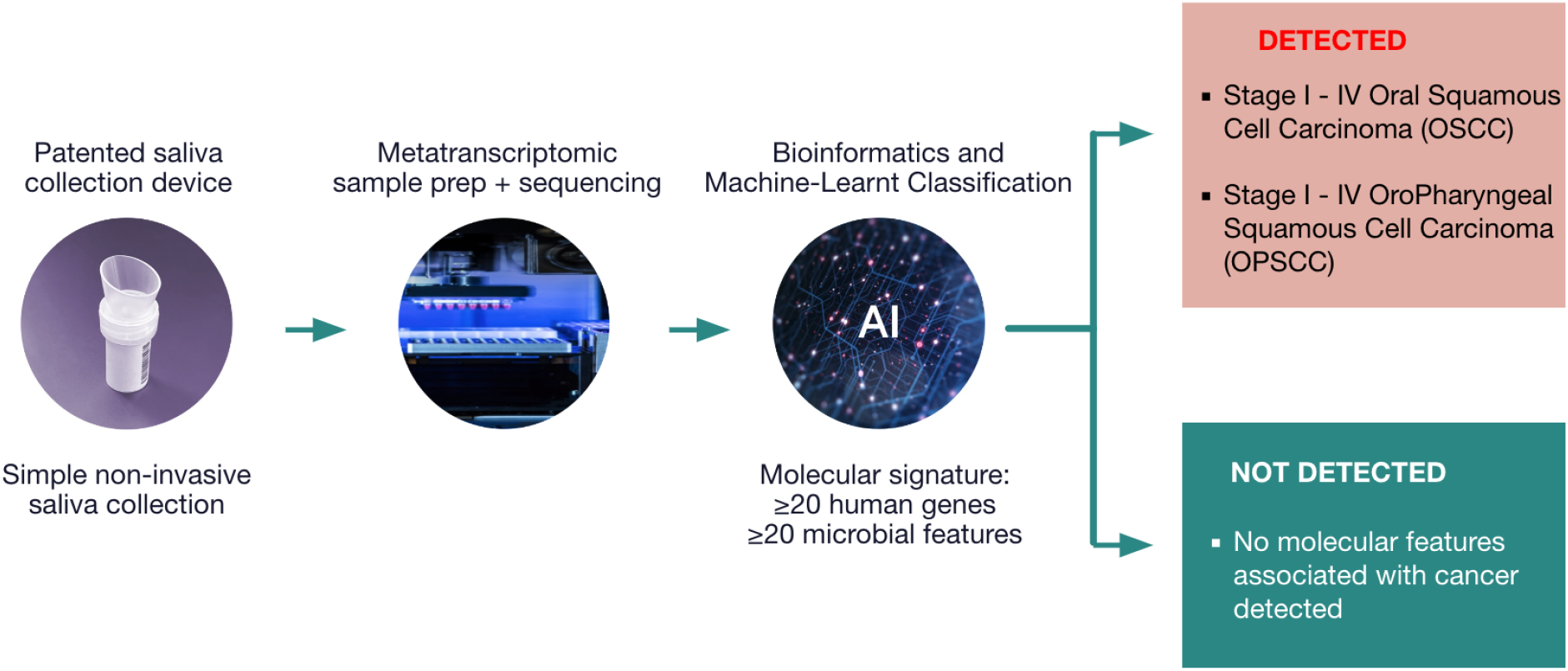
Overview of salivary RNA metatranscriptomic signature based cancer detection system.

### (i) Sample collection and transport

Unstimulated whole mouth saliva samples were collected as published previously [23]. The test includes a custom collection device, which allows easy collection of a saliva sample at home by individuals, or during oral examination by a qualified healthcare professional. The collection tube contains a preservative that dissolves cell membranes and penetrates all cells, denatures nucleases, and prevents RNA self-cleaving by preventing deprotonation of the 2’-OH. The use of this proprietary preservative enables ambient temperature transportation for saliva samples. Saliva sample collection, preservation, transportation and lab preparation are described in Banavar et al. [23].

### (ii) Lab processing

Our CLIA-certified lab receives the saliva samples and processes it to extract and sequence the RNA from the saliva sample. Our test extracts and sequences all mRNA molecules in a non-discriminatory fashion, after eliminating the non-informative rRNA molecules. After sample preparation is completed in the Lab, total RNA is extracted from clarified lysate using a custom silica bead-based protocol, which includes on-bead DNA removal by DNase. Total RNA is quantified using the RiboGreen method and diluted when necessary. Bacterial and human rRNAs are physically removed from the specimen using a subtractive hybridization method. The remaining RNAs are converted into Illumina directional sequencing libraries [24]. Library pools are then sequenced on Illumina NovaSeq 6000 to produce sequencing data.

### (iii) Data processing

The sequenced data is processed through our bioinformatics pipeline and an OSCC/OPSCC classifier. The bioinformatics pipeline maps sequenced reads to human genes (or HG), as well as microbial species (or SP) and microbial gene clusters annotated as KEGG Orthologs (or KO) [23]. For HG detection, paired-end reads are mapped to the human transcriptome. Gene expression levels are computed by collecting the transcript-level abundance (transcript per million - TPM) and then aggregating them to the gene level using Salmon version 1.1.0 [25]. For taxonomic classification, reads are mapped to a custom catalog derived from genomic sequences from all domains of the phylogenetic tree, namely, bacteria, archaea, eukaryota, and viruses. Taxonomies are identified and their relative activities are calculated at three different taxonomic ranks (genus, species, and strain). To identify and quantify transcriptionally active genes in the microbial community, functional assignments are obtained through alignment of the sequencing reads to another custom curated catalog of genes and the KEGG databases [26]. Further details of the bioinformatics processing can be found in Banavar et al. [23]. All the detected molecular features (HG, SP, and KO features) are then used for downstream analyses including classifier development, validation, and eventually, classification of new samples.

The OSCC/OPSCC classifier is a machine-learning (ML) model that uses the HG, SP, and KO features and classifies the sample as belonging to the “OSCC/OPSCC class” or the “Not OSCC/OPSCC class” within pre-specified performance criteria. The overall workflow of classification model development and independent validation is shown in Figure 2. Model development is described in detail in the supplementary materials. The final trained model coming out of the development phase was determined to be capable of inferring, at a high probability, whether a participant’s sample has OSCC and/or OPSCC or not, and used for independent validation with unseen samples, on the other side of the “firewall” in Figure 2.

**Figure 2.**
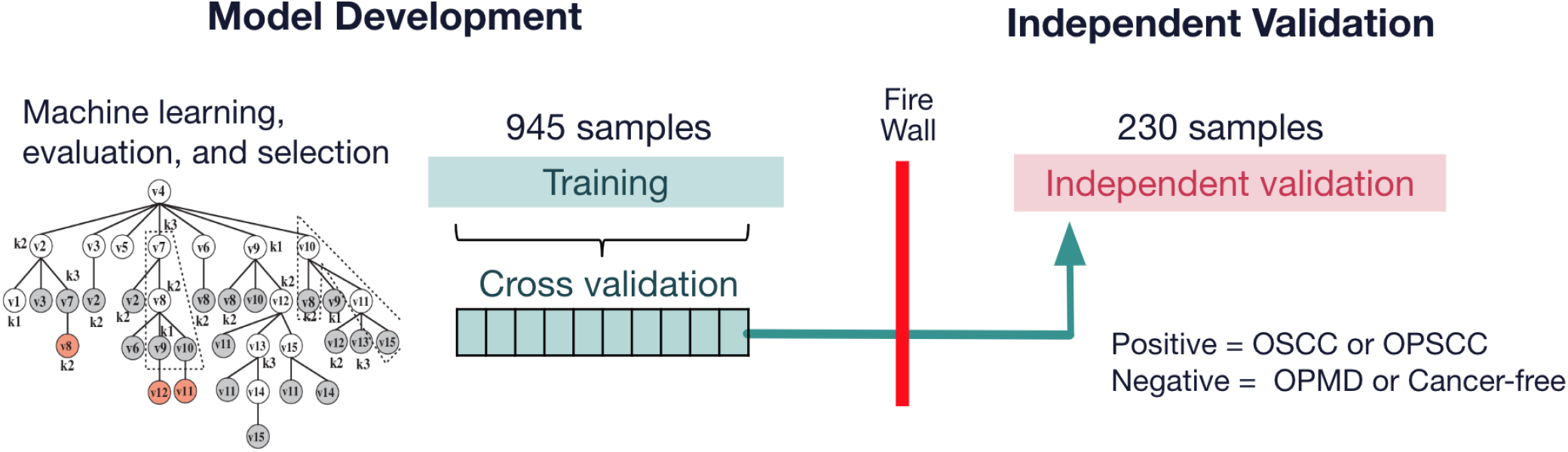
Model development, cross validation and independent validation workflow

## Patent Population

The CDOT test was developed with samples from a cohort of 945 individuals (details in the supplementary materials), and validated in an unseen independent cohort of 230 individuals in order to evaluate its performance in the proposed intended use. In this section, we describe the patient population on which the test was validated.

Validation study participants were either 50 years or older or had a history of tobacco use. Having a “history of tobacco use” included being a current or former tobacco user per the Affordable Care Act (ACA) definition. A current tobacco user was defined as someone who uses tobacco products four or more times per week in the past six months. A former tobacco user is a person who has quit using tobacco products at the current time but had previously used tobacco products four or more times per week for six months or more, within the last 20 years. For purposes of this study, we defined tobacco broadly consistent with definitions for multiple health organizations, including the World Health Organization (WHO), which defines “tobacco use” to include smoking, sucking, chewing or snuffing any tobacco product.

The validation cohort of 230 included 101 samples from secondary care centers, and a combination of 129 clinically adjudicated patients with OPMD and cancer free patients from the Viome company customer database in the US (Table 2). Eligible participants had to be free from any active infection, have no cancer in the past, not be pregnant and have no irradiation to the neck and head region. The study was approved by the Queensland University of Technology and University of Queensland Medical Ethical Institutional Boards (HREC no.: 1400000617 and HREC no.: 2017000662 respectively) and the Royal Brisbane and Women’s Hospital (HREC no.: HREC/12/QPAH/381) Ethics Review Board. All participants gave their consent to participate in the study.

**Table 2.** Independent validation cohort (n= 230) OSCC, Oral squamous cell carcinoma; OPSCC, Oropharyngeal squamous cell carcinoma; OPMD, Oral premalignant disorder

|  |  |  |  |  |
| --- | --- | --- | --- | --- |
|  | Positives (n= 96) |  | Negatives (n=134) |  |
|  | OSCC (n= 20) | OPSCC (n= 76) | OPMD (n=14) | Cancer free (n= 120) |
| **Female (n)** | 3 (15.0%) | 4 (5.3%) | 7 (50.0%) | 42 (35.0%) |
| **Age (yrs) mean ± SD** | 61.3 ± 11.1 | 60.9 ± 7.6 | 62.9 ± 15.9 | 60.5 ± 8.8 |

Patients with OSCC or OPSCC were clinically diagnosed to confirm their cancer status. Clinical data also included histopathology reports after biopsying the patients, spanning early (Stage I/II) and late (Stage III/IV) stage OSCC and OPSCC. OSCC and OPC diagnosis was performed with biopsy and examination of FFPE tissue sections by routine (Hematoxillin and eosin) stain using standard methodology. Pathological Staging of OSCC and OPC was also performed. Largest diameter of tumor, tumor thickness and presence of bone invasion are essential for T stage categorization. Lymph node stage is mainly based on the number and size of involved lymph nodes, laterality of involved nodes, and presence or absence of extranodal extension of tumor deposit.

Patients with oral premalignant disorders (OPMD) or cancer-free participants could be clinically adjudicated by a primary physician. OPMD included the following conditions: dysplasia, hyperplasia, leukoplakia, erythroplakia, lichenoid lesions, actinic keratosis and lichenoid reaction; as well as canker sores, gingival enlargement as a result of a dental procedure, lichen planus, keratosis, inflammatory reaction and cheek bites.

## Results

We evaluated the performance of the CDOT test using various metrics. Each participant sample was analyzed using the classifier and the results were compared to the participant’s known or assumed (cancer-free volunteers were assumed to be cancer-free) cancer status to determine the classifier’s performance characteristics (Figure 2). Specificity and sensitivity were also evaluated by disease stage (early vs late), smoking status (current, former, non-smoker and unknown) and age (<50 and >=50).

Figure 3a and 3b show the area under the receiver operating characteristic (ROC) curve (ROC - AUC) and the distributions of the predicted probabilities for the model on the independent validation data set. The AUC is 96%, indicating a probability of 0.96 that our classifier will rank a randomly chosen positive instance higher than a randomly chosen negative one (assuming ‘positive’ ranks higher than ‘negative’).

**Figure 3a:**
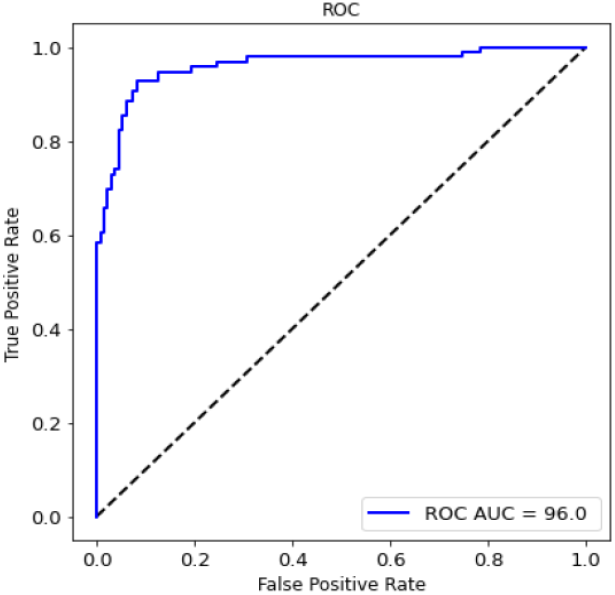
ROC plot for the model on independent validation set

**Figure 3b:**
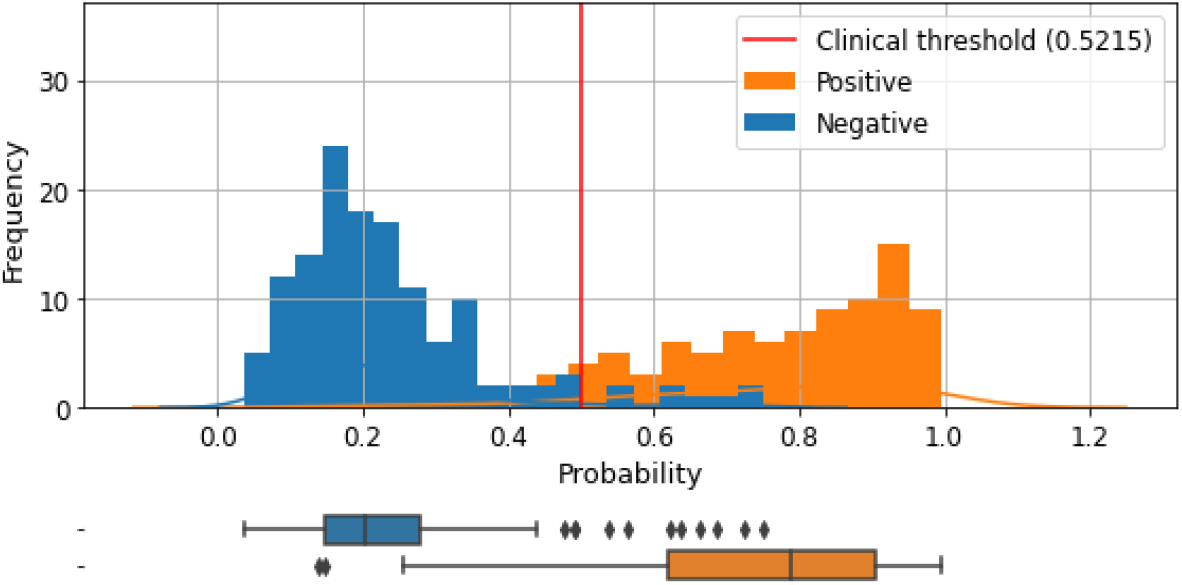
Predicted probabilities for all participants in the independent validation set

Further, the OSCC-OPSCC classifier correctly classified 18/20 = 90% OSCC positive patients (sensitivity to OSCC), 64/76 = 84.2% OPSCC positive patients (sensitivity to OPSCC) (Table 3a) and 126/134 = 94% negative participants as no cancer (specificity no negative samples) (Table 3b). Out of the early stage participants with OSCC or OPSCC, the OSCC-OPSCC classifier was able to classify 9 out of the 10 as OSCC and 51 out of 62 as OPSCC positive thereby demonstrating a reasonable expectation of clinical success in identifying participants with OSCC and/or OPSCC, including those with early-stage disease (Table 3a). Also, in Table 3b, we included the breakdown of the model’s specificity to negative samples.

**Table 3a:** Sensitivity (Positive percent agreement) OSCC, Oral squamous cell carcinoma; OPSCC, Oropharyngeal squamous cell carcinoma

|  | n/N (%) |
| --- | --- |
| Overall sensitivity (TP / TP + FN) | 82/96 (85.0%)<br>95% CI [76.7%, 91.8%] |
| OSCC sensitivity | 18/20 (90.0%) |
| OSCC Early stage | 9/10 (90.0%) |
| OSCC Late stage | 9/10 (90.0%) |
| OPSCC sensitivity | 64/76 (84.2%) |
| OPSCC Early stage | 51/62 (82.3%) |
| OPSCC Late stage | 13/14 (92.9%) |

**Table 3b:**
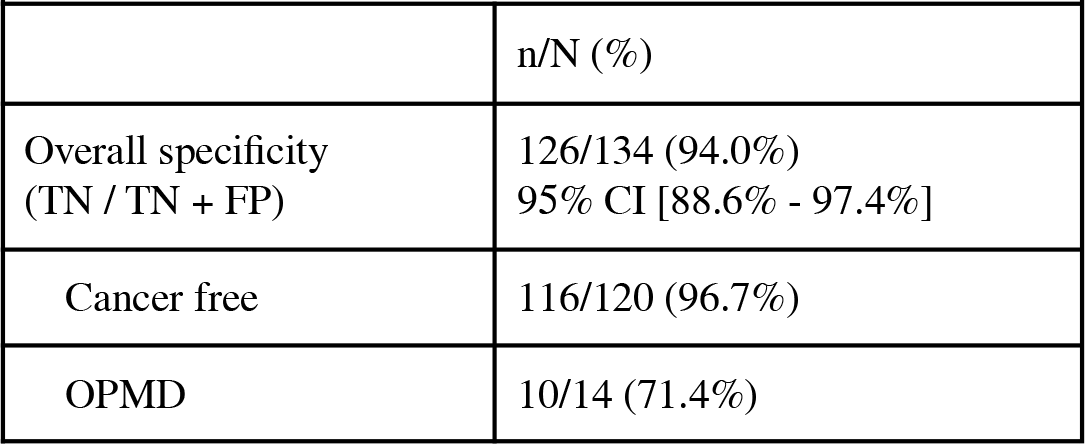
Specificity (Negative percent agreement) OPMD, Oral Premalignant Disorder

|  | n/N (%) |
| --- | --- |
| Overall specificity (TN / TN + FP) | 126/134 (94.0%)<br>95% CI [88.6% - 97.4%] |
| Cancer free | 116/120 (96.7%) |
| OPMD | 10/14 (71.4%) |

We stratified the patient characteristics across the care centers used to source the patient samples for validation, to ensure that there was no bias. When evaluating the performance of the model by smoking status, the OSCC-OPSCC classifier correctly classified 100% of the current smokers. Among former smokers, 7/8 = 87.5% OSCC and 35/41 (85.4%) OPSCC were correctly classified as positives. Among non-smokers, 4/4 = 100% OSCC and 13/17 (76.5%) OPSCC were correctly classified as positives (Table 3c).

**Table 3c:** Sensitivity and specificity by smoking status OSCC, Oral squamous cell carcinoma; OPSCC, Oropharyngeal squamous cell carcinoma

|  | Sensitivity (n/N) % |  | Specificity (n/N) % |
| --- | --- | --- | --- |
|  | OSCC | OPSCC |  |
| Current | 6/6 (100%) | 11/11 (100%) | 2/4 (50%) |
| Former | 7/8 (87.5%) | 35/41 (85.4%) | 5/7 (71.4%) |
| Non-smoker | 4/4 (100%) | 13/17 (76.5%) | 117/121 (97.7%) |
| Unknown | 1/2 (50%) | 5/7 (71.4%) | 2/2 (100%) |

When evaluating the performance of the model by age, among people below 50 years old, 4/4 = 100% of people with OSCC and 2/3 = 66.7% of people with OPSCC were correctly classified as positives. Among older people, 15/17 = 88.2% OSCC and 62/73 (84.9%) OPSCC were correctly classified as positives (Table 3d). Similarly, when stratifying the data by biological sex, we observed that the distribution of positive and negative samples across disease state was concordance between male and female.

**Table 3d:** Sensitivity and specificity by age group OSCC, Oral squamous cell carcinoma; OPSCC, Oropharyngeal squamous cell carcinoma

|  | Sensitivity (n/N) % |  | Specificity (n/N) % |
| --- | --- | --- | --- |
|  | OSCC | OPSCC |  |
| < 50 years | 3/3 (100%) | 2/3 (66.7%) | 4/5 (80%) |
| >= 50 years | 15/17 (88.2%) | 62/73 (84.9%) | 122/129 (94.5%) |

An interference evaluation was also performed with 41 cancer free negative participants. Participants were required to chew gum, chew tobacco, and brush their teeth. These analyses determined whether external interference factors influenced the detection power of the model. The probability output of the model did not change based on the presence of the different interfering substances, showing the robustness of the model to interferants (Supplementary Figure 2).

## Discussion

This study evaluates the effectiveness of a saliva metatranscriptomic detection test, CDOT™, to identify individuals with OSCC and OPSCC. To evaluate its performance, the test result (negative or positive) was compared with the histopathological diagnosis of all significant lesions discovered during a biopsy. Based on this comparison, CDOT™ sensitivity (true positive fraction) is 90% for participants with a histopathological diagnosis of OSCC and 84.2% for participants with a diagnosis of OSPCC. The test can also detect true positives in early and late stages of OSCC with 90% sensitivity. Furthermore, among participants having a valid oral cancer test result and a self-reported or clinically adjudicated cancer free status, specificity for cancer free patients not including OPMD is 96.7%. The specificity or ability of the test to designate true negatives is 94%, bringing a new paradigm for early screening in primary care settings and reducing the number of unnecessary biopsies in secondary care settings.

In routine clinical practice today, the diagnostic pathway for oral cancer is dependent on the experience and expertise of different healthcare providers, including dentists, dental hygienists and primary physicians who are responsible for performing the head and neck visual examinations. Oral lesions that may be indicative of oral cancer include heterogeneous appearance such as changes in color, texture and size; and alterations in the surface, for example, non-healing ulcerations. Several adjunct diagnostic tools are available to aid providers in identification and diagnosis, but there is no general consensus on which, if any, is most reliable. Examples are exfoliative cytology, including liquid-based, scraped and brush cytology [27,28], toluidine blue staining [29], and light-based visual detection systems [30]. The performance of these methods vary widely, with a pooled estimate of 88% sensitivity and 81% specificity [31], which is a lower performance when compared to the proposed saliva-based detection test. Furthermore, CDOT™ is non-invasive and easy to use and transport, which makes it a good screening tool in dentist and primary physician’s offices.

With the advent of AI technologies there are many new imaging methods introduced in the last decade that act as adjunctive technologies, ranging from fluorescent imaging to optical and mobile phone imaging in teledentistry [32] [33]. Fluorescent imaging is a non-invasive method supported with confocal laser endomicroscopy which has high magnification power [34] resulting in 92% specificity, as well as the N2 laser study with 92% specificity [35]. These laser technologies are still expensive and not readily available for primary care settings. In addition, there is significant upskilling required for thorough examination and the results might be operator dependent. Another example is the Oncogrid surveillance program [36] which uses mobile phones connecting primary care dental practitioners and frontline health workers with oral cancer specialists for screening oral cancer. While these methods are easily accessible, they may only be partially useful for certain low resource setting areas, as they have low sensitivity (around 70%-85%) due to the limitations of the access to certain areas of the mouth cavity [37] [38]. In contrast, the advantage of saliva-based methods is that they can be easily accessible in both primary and secondary care settings, and the use of technology is not operator dependent.

Saliva is in direct contact with the tissues of the oral cavity and represents a biofluid which acts as a great substrate for liquid biopsy. The biomolecules detected by our metatranscriptomic method offer deep resolution and insight into the activity of the human genes as well as all the microbial species – making it the first of its kind. Furthermore, our method uses advanced machine learning modeling to tease out the most distinguishing molecular features associated with OSCC and/or OPSCC. Previous methods have either assessed biomolecules from the human side (e.g. CD44 protein [39] or RNA – 6 markers that include interleukins IL-1Beta, IL-8, OAZ1SAT1S100P, and DUSP1 [40] or difference in species with 16S rRNA gene sequencing and metagenomics [41] [42], but their results are less promising, and while they have reported discovery results, they have not been validated in independent cohorts. Large independent prospective studies are required to advance adoption of the new adjunctive technologies in clinical and eventually in home-based testing settings.

If a secondary care specialist has a suspicion of cancer, the patient will undergo a biopsy, a common invasive procedure that remains the gold standard for diagnosing premalignant and malignant oral diseases. CDOT™ is non-invasive and can be easily included in secondary care practices to confirm the need for a biopsy. Oral biopsy involves both psychological implications for the patient and technical difficulties for the health practitioner. When lesions are extensive, the most representative areas must be selected to avoid diagnostic errors. In fact, inter- and intra-observer variability of histological diagnosis for dysplasia is well documented [43].

A useful diagnostic tool should be easy to use and cause minimal patient discomfort. Ideally, a diagnostic procedure should be neither time-consuming nor complicated and, in addition to high sensitivity, should have the potential for automation. High specificity also avoids false-positives and, therefore, reduces patient anxiety, additional investigations, and even unnecessary treatment. This precisely describes the oral CDOT™ test. CDOT™ provides non-invasive information regarding a patient’s OSCC or OPSCC disease status that can aid in seeking a definitive diagnosis and treatment planning. It offers significant advantages over existing alternatives because of its high sensitivity and specificity, and it has the potential to identify patients for additional follow-up before their disease has progressed to be apparent in visual/tactile exams (i.e., Stages I/II).

We estimate the prevalence of Oral Cancer in the United States at 0.4%. With this prevalence, the Positive Predictive Value (PPV) for our test is 5.4%, including both OSCC and OPC in the intended use. The corresponding Negative Predictive Value (NPV) for our test is 99.9%. Also, the positive likelihood ratio (LR+) of our test is 14.31 and the negative likelihood ratio (LR-) is 0.16. Since the inverse of negative likelihood ratio (6.25) is less than LR+, based on these data, we conclude that our test will be used as a rule-in test.

An important goal of early detection of oral cancer is to shift from detection at Stage III/IV to detection at Stage I/II. To demonstrate this stage-shifting benefit, we developed a cancer intercept micro-simulation model to evaluate the use of the Viome device (point estimate sensitivity of 90%, lower 95% confidence limit of 68%) within a large intended use population, with the distribution of oral cavity and pharynx cancers using data from SEER (https://seer.cancer.gov/), a large validated database. In one simulation, a cohort of 30-year-olds was generated, and cancers were allowed to develop up to age 50. At age 50, a one-time screen is applied, and the cohort is followed from age 50-53 (3 year follow-up). Cancer stage distribution is evaluated from cancers that are detected (from the screen) and those that present with clinical symptoms prior to age 53. All detected cancers are treated with the standard of care. SEER-derived age-specific cancer incidence rates are used. The simulated one-time screening strategy is then compared to no screening, where an identical population is simulated, and cancer stage distribution is aggregated from age 50-53. In this simulation, with a 5-year sojourn time (length of time the cancer remains asymptomatic), we found that the proportion of early and late stage oral cancers in the no-screening scenario was 29% and 71% respectively, whereas in the Viome one-time screening scenario it was 56% and 44% respectively. This illustrates that 27% of late-stage cases were stage-shifted to the early stage, thereby enabling a standard of care treatment for those cases and concomitant benefits.

We recognize that there are some limitations to our study. While the model performs well in patients with OSCC and OPSCC and cancer free patients, the number of participants with pre-malignant diseases is currently too low to discriminate between positives and negatives. The model was validated in former and current smokers as well as young and older participants, however other populations at risk such as heavy drinkers and patients with HPV-OPSCC were not evaluated and deserve further exploration. Lastly, while the study participants were recruited from across the US and Australia, the positive cases were recruited from a single site in Australia. A larger multi-site validation including centers in the US is forthcoming, and is expected to address most of these limitations.

In summary, CDOT™ is a saliva-based detection test for oral cavity cancers and oropharyngeal cancers, with 94% specificity and sensitivity of 90% for OSCC and 84% for OPSCC. The test performs RNA sequencing analysis and uses 270 human and microbial mRNA features as markers associated with oral and throat cancer. Our machine-learning based test was validated on an independent cohort of 230 patients. While future studies with a larger number of patients with pre malignancies are needed, the current method is a practically useful, non-invasive method that can be easily incorporated in dentist offices, primary care centers and specialized cancer clinics for early detection of oral and throat cancers.

## Data Availability

Data produced in the present study are available upon reasonable request to the authors

https://www.viomelifesciences.com/data-access

## Supplementary Materials

### Classification model development

As shown in Supplementary Table 1, the ML training set consisted of samples from 945 participants, with 92 samples labeled as positives (80 OSCC and 12 OPSCC) and 853 samples labeled as negatives (805 cancer free participants, and 48 with pre-malignant disorders). Out of the 945 training samples, 744 came from primary care centers as well as Viome customers distributed across the US, and 201 from a secondary care hospital center and cancer-free individuals in Australia.

An OSCC-OPSCC binary classifier, a machine-learning model, was built using a *l*_*2*_ regularized logistic regression algorithm after evaluating several model candidates. An internal k-fold cross-validation (CV) was performed within the 945 samples. Model hyperparameters included the number of folds (k), variance threshold (v), regularization strength (C), p-value for selecting stable features via bootstrapping (p) and clinical threshold (t) that maximized the sum of sensitivity and specificity. A total of 93 hyperparameter sets (models) reached at least 90% specificity and sensitivity across the cross-validation, and were further inspected for ROC-AUC, sensitivity, specificity and the variance of the performance metrics. We selected the model that had the highest performance score, defined as the sum of average CV sensitivity and average CV specificity, among the models trained on a feature set containing human genes. This best model had k, v, C, p and t to be 5, 25, 0.01, 0.1 and 0.5215, respectively. Then the model with the best hyperparameters (from the CV step) was trained with the entire 945 samples to obtain the final/frozen machine-learning model (Suppl. Table 1). The final model encapsulated a pattern of molecular data that correlated with OSCC and OPSCC, which is referred to as the “molecular signature” of OSCC-OPSCC. The molecular signature consisted of 270 features: 110 species, 72 microbial gene clusters annotated as KO’s and 88 HGs. A full biological interpretation of these molecular features is outside the scope of this manuscript, but we summarize the features in Supplementary Figure 1a, which aggregates the 182 microbial features into functional categories, and Supplementary Figure 1b which aggregates the 88 human gene features into known cancer hallmarks.

**Supplementary table 1.** Model development (training) cohort (n=945) OSCC, Oral squamous cell carcinoma; OPSCC, Oropharyngeal squamous cell carcinoma; OPMD, Oral premalignant disorder.

|  | Positives (n=92) |  | Negatives (n=853) |  |
| --- | --- | --- | --- | --- |
|  | OSCC<br>(n= 80) | OPSCC<br>(n= 12) | OPMD<br>(n=48) | Cancer free<br>(n= 805) |
| **Female (n)** | 23 (28.8%) | 0 (0.0%) | 29 (60.4%) | 561 (69.7%) |
| **Age (yrs) mean ± SD** | 59.4 ± 13.9 | 59.8 ± 8.8 | 61.5 ± 11.4 | 44.0 ± 13.7 |

**Supplementary Figure 1a.**
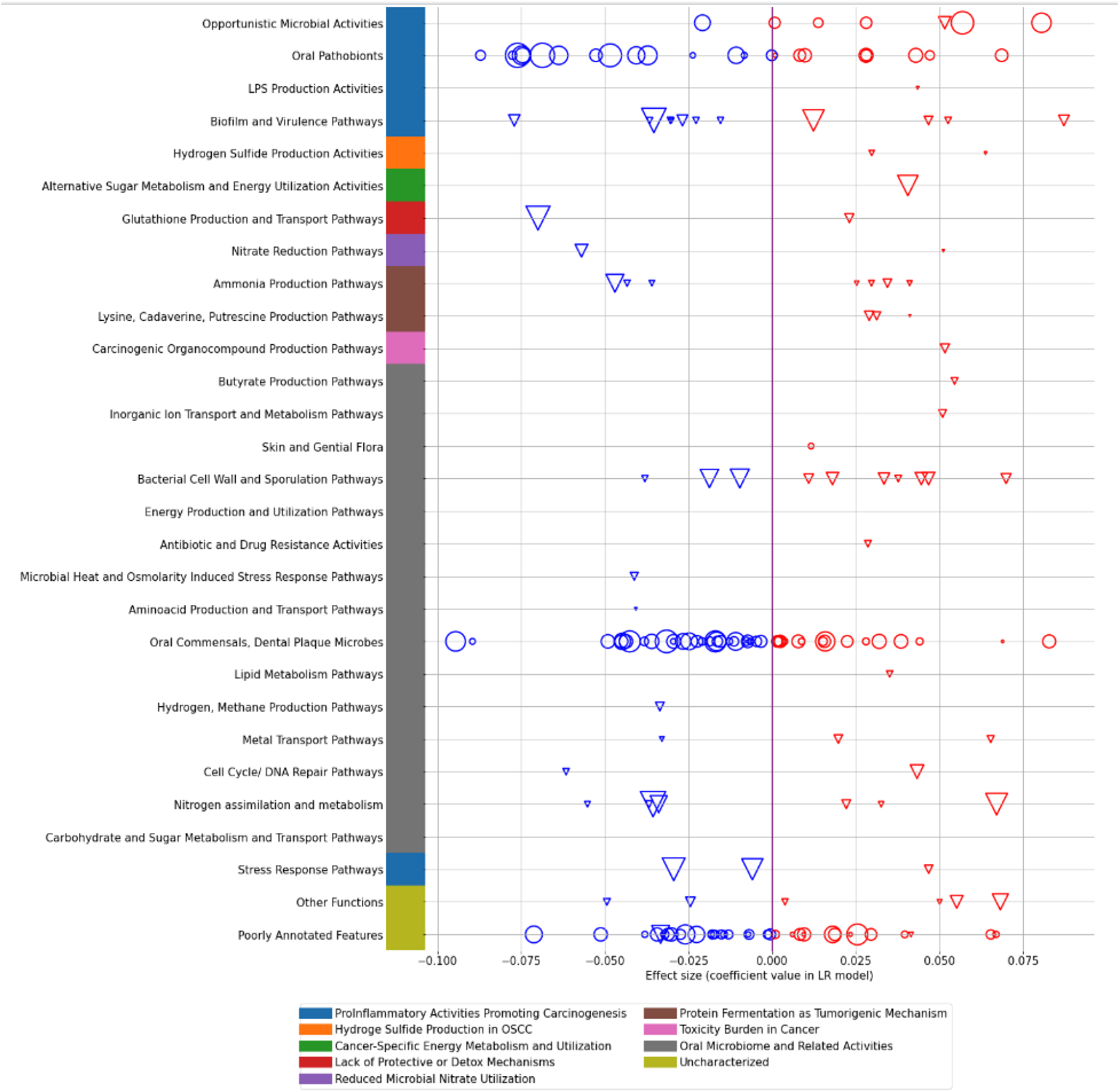
182 microbial features (110 species and 72 KOs) in the molecular signature of the final machine-learnt model. Circles and triangles represent active species and active KOs respectively. Sizes of circles or triangles are proportional to the CLR median difference in expression level between cases and controls. Features with coefficients greater than zero (shown in red) denote higher expression in cancer cases, and those with coefficients less than or equal to zero (shown in blue) denote higher expression in cancer negatives. LPS is lipopolysaccharides. These microbial features were curated and aggregated into functional categories by systems biology experts using the available literature relating to each of the features.

**Supplementary Figure 1b.**
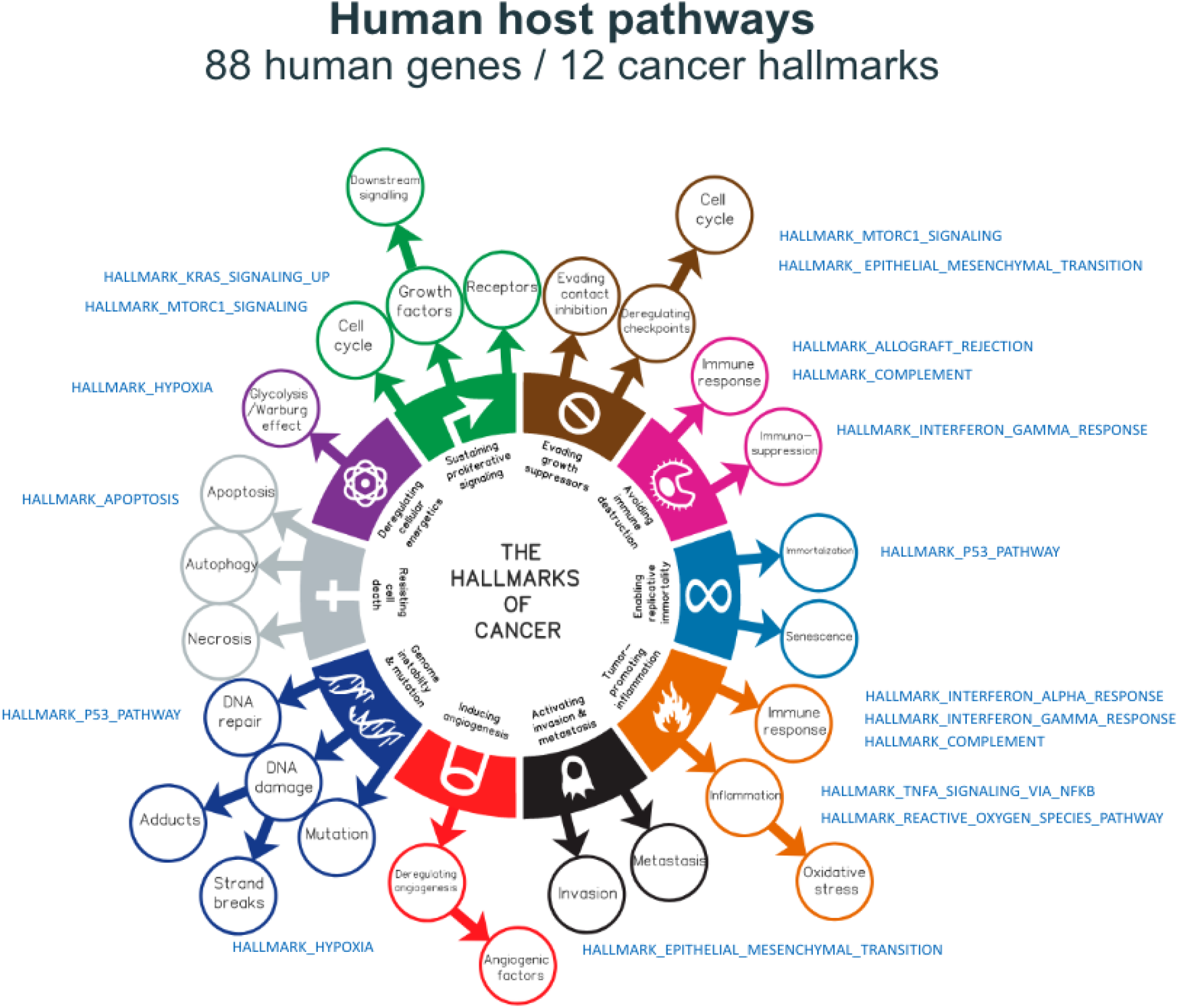
88 human gene features in the molecular signature of the final machine-learnt model. The 88 human genes have a statistically significant overlap with several cancer hallmark genesets such as interferon Gamma, interferon Alpha, KRAS signaling and p53 pathways, with an analysis done via a Gene Set Enrichment Analysis (GSEA) tool. GSEA computes overlaps with a Molecular Signatures Database (MSigDB), a collection of annotated gene sets divided into major collections, representing a universe of biological processes and pathways which are meaningful for insightful interpretation, each based on published experimental findings. This analysis shows that the 88 human gene features in our model represent known associations with the biology of cancer.

**Supplementary Figure 2:**
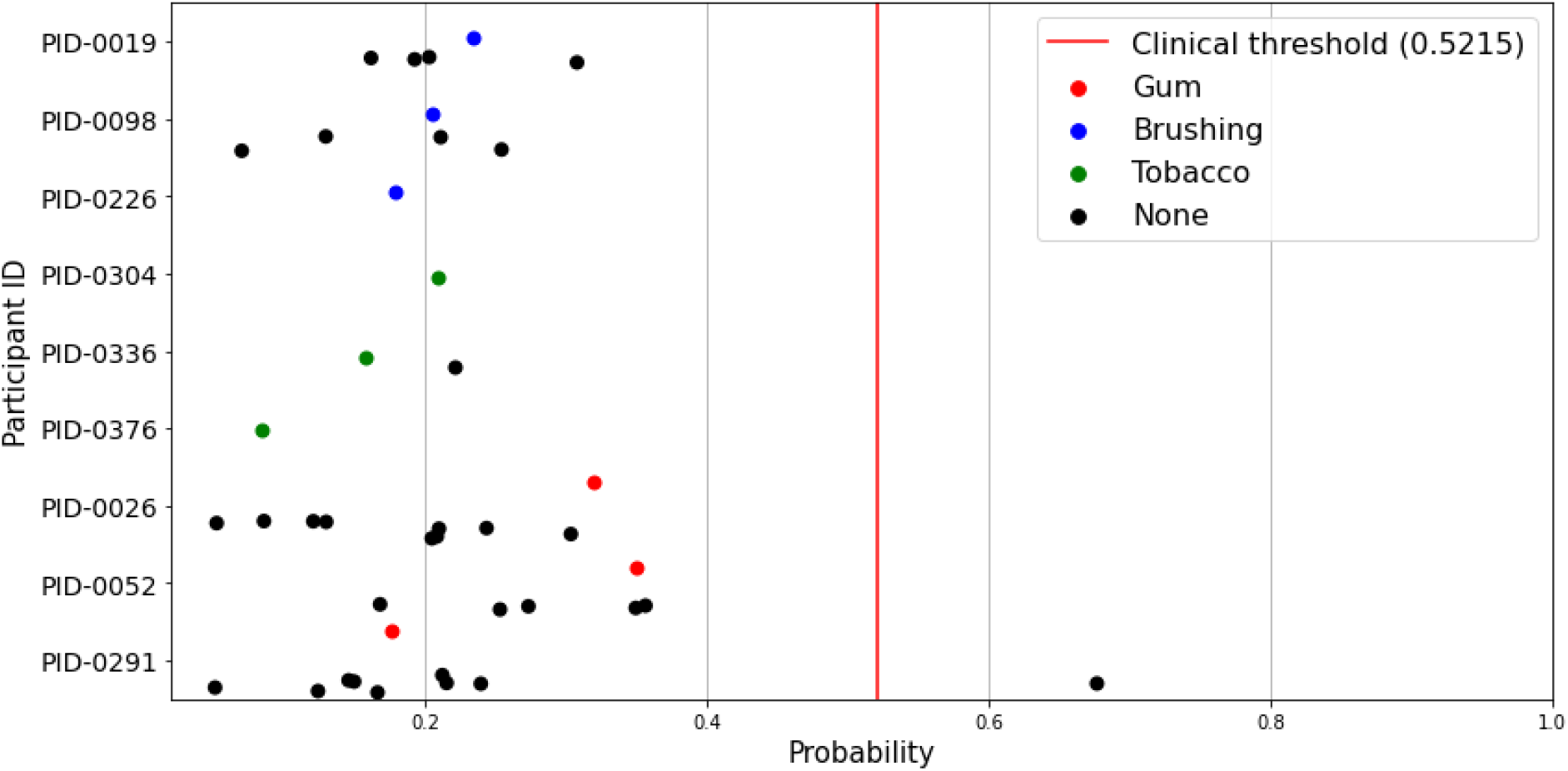
Model predicted probabilities for interference samples. The test was evaluated for various forms of interference experienced shortly before the collection of the saliva samples, namely: gum, brushing and tobacco and no interference (none). For this experiment, 41 independent samples were collected, processed and evaluated, from cancer-free individuals that had experienced interference prior to the collection of saliva samples. The result highlights the stability of the test result, even when there is interference before sample collection.

## References

[1] Bosetti C, Carioli G, Santucci C, Bertuccio P, Gallus S, Garavello W, et al. Global trends in oral and pharyngeal cancer incidence and mortality. Int J Cancer 2020;147:1040–9. https://doi.org/10.1002/ijc.32871.

[2] Key statistics for oral cavity and oropharyngeal cancers. 2022.

[3] Cheong SC, Vatanasapt P, Yi-Hsin Y, Zain RB, Kerr AR, Johnson NW. Oral cancer in South East Asia. Transl Res Oral Oncol 2017;2:2057178X17702921. https://doi.org/10.1177/2057178x17702921.

[4] Joshi P, Dutta S, Chaturvedi P, Nair S. Head and Neck Cancers in Developing Countries. Rambam Maimonides Medical J 2014;5:e0009. https://doi.org/10.5041/rmmj.10143.

[5] Weeramange CE, Liu Z, Hartel G, Li Y, Vasani S, Langton-Lockton J, et al. Salivary High-Risk Human Papillomavirus (HPV) DNA as a Biomarker for HPV-Driven Head and Neck Cancers. J Mol Diagnostics 2021;23:1334–42. https://doi.org/10.1016/j.jmoldx.2021.07.005.

[6] Lim Y, Tang KD, Karpe AV, Beale DJ, Totsika M, Kenny L, et al. Chemoradiation therapy changes oral microbiome and metabolomic profiles in patients with oral cavity cancer and oropharyngeal cancer. Head Neck 2021;43:1521–34. https://doi.org/10.1002/hed.26619.

[7] Tang KD, Vasani S, Menezes L, Taheri T, Walsh LJ, Hughes BGM, et al. Oral HPV16 DNA as a screening tool to detect early oropharyngeal squamous cell carcinoma. Cancer Sci 2020;111:3854–61. https://doi.org/10.1111/cas.14585.

[8] Cristaldi M, Mauceri R, Fede OD, Giuliana G, Campisi G, Panzarella V. Salivary Biomarkers for Oral Squamous Cell Carcinoma Diagnosis and Follow-Up: Current Status and Perspectives. Front Physiol 2019;10:1476. https://doi.org/10.3389/fphys.2019.01476.

[9] Peacock ZS, Pogrel MA, Schmidt BL. Exploring the Reasons for Delay in Treatment of Oral Cancer. J Am Dent Assoc 2008;139:1346–52. https://doi.org/10.14219/jada.archive.2008.0046.

[10] Warnakulasuriya S, Kerr AR. Oral Cancer Screening: Past, Present, and Future. J Dent Res 2021;100:1313–20. https://doi.org/10.1177/00220345211014795.

[11] LeHew CW, Epstein JB, Kaste LM, Choi Y. Assessing oral cancer early detection: clarifying dentists’ practices. J Public Health Dent 2010;70:93–100. https://doi.org/10.1111/j.1752-7325.2009.00148.x.

[12] ADA Expands Policy On Oral Cancer Detection To Include Oropharyngeal Cancer, Oral Health Group (Oct. 3, 2019). n.d.

[13] Centers for Disease Control and Prevention. Quickstats: percentage of adults aged >18 years who have ever had an oral cancer examination, by smoking status and age group. United States: National Health Interview Survey; 2008.

[14] Chhabra N, Chhabra S, Sapra N. Diagnostic Modalities for Squamous Cell Carcinoma: An Extensive Review of Literature-Considering Toluidine Blue as a Useful Adjunct. J Maxillofac Oral Surg 2015;14:188–200. https://doi.org/10.1007/s12663-014-0660-6.

[15] Shyamala K, Girish HC, Murgod S. Risk of tumor cell seeding through biopsy and aspiration cytology. J Int Soc Prev Community Dent 2014;4:5–11. https://doi.org/10.4103/2231-0762.129446.

[16] Vimal J, Himal I, Kannan S. Role of microbial dysbiosis in carcinogenesis & cancer therapies. Indian J Medical Res 2020;152:553–61. https://doi.org/10.4103/ijmr.ijmr_1026_18.

[17] Hajishengallis G, Liang S, Payne MA, Hashim A, Jotwani R, Eskan MA, et al. Low-Abundance Biofilm Species Orchestrates Inflammatory Periodontal Disease through the Commensal Microbiota and Complement. Cell Host Microbe 2011;10:497–506. https://doi.org/10.1016/j.chom.2011.10.006.

[18] Chen C, Hemme C, Beleno J, Shi ZJ, Ning D, Qin Y, et al. Oral microbiota of periodontal health and disease and their changes after nonsurgical periodontal therapy. Isme J 2018;12:1210–24. https://doi.org/10.1038/s41396-017-0037-1.

[19] Mager D, Haffajee A, Devlin P, Norris C, Posner M, Goodson J. The salivary microbiota as a diagnostic indicator of oral cancer: a descriptive, non-randomized study of cancer-free and oral squamous cell carcinoma subjects. J Transl Med 2005;3:27. https://doi.org/10.1186/1479-5876-3-27.

[20] Su S-C, Chang L-C, Huang H-D, Peng C-Y, Chuang C-Y, Chen Y-T, et al. Oral microbial dysbiosis and its performance in predicting oral cancer. Carcinogenesis 2020;42:127–35. https://doi.org/10.1093/carcin/bgaa062.

[21] Wang L, Yin G, Guo Y, Zhao Y, Zhao M, Lai Y, et al. Variations in Oral Microbiota Composition Are Associated With a Risk of Throat Cancer. Front Cell Infect Mi 2019;9:205. https://doi.org/10.3389/fcimb.2019.00205.

[22] M. P, Al-hebshi N, Perera I, Ipe D, Ulett G, Speicher D, et al. Inflammatory Bacteriome and Oral Squamous Cell Carcinoma. J Dent Res 2018;97:725–32. https://doi.org/10.1177/0022034518767118.

[23] Banavar G, Ogundijo O, Toma R, Rajagopal S, Lim YK, Tang K, et al. The salivary metatranscriptome as an accurate diagnostic indicator of oral cancer. Npj Genom Medicine 2021;6:105. https://doi.org/10.1038/s41525-021-00257-x.

[24] Hatch A, Horne J, Toma R, Twibell BL, Somerville KM, Pelle B, et al. A Robust Metatranscriptomic Technology for Population-Scale Studies of Diet, Gut Microbiome, and Human Health. Int J Genomics 2019;2019:1718741. https://doi.org/10.1155/2019/1718741.

[25] Patro R, Duggal G, Love MI, Irizarry RA, Kingsford C. Salmon provides fast and bias-aware quantification of transcript expression. Nat Methods 2017;14:417–9. https://doi.org/10.1038/nmeth.4197.

[26] Almeida A, Nayfach S, Boland M, Strozzi F, Beracochea M, Shi ZJ, et al. A unified catalog of 204,938 reference genomes from the human gut microbiome. Nat Biotechnol 2021;39:105–14. https://doi.org/10.1038/s41587-020-0603-3.

[27] Charanya D, Raghupathy LP, Farzana AF, Murugan R, Krishnaraj R, Kalarani G. Adjunctive aids for the detection of oral premalignancy. J Pharm Bioallied Sci 2016;8:S13–9. https://doi.org/10.4103/0975-7406.191942.

[28] Alsarraf AH, Kujan O, Farah CS. The utility of oral brush cytology in the early detection of oral cancer and oral potentially malignant disorders: A systematic review. J Oral Pathol Med 2018;47:104–16. https://doi.org/10.1111/jop.12660.

[29] Pallagatti S, Sheikh S, Aggarwal A, Gupta D, Singh R, Handa R, et al. Toluidine blue staining as an adjunctive tool for early diagnosis of dysplastic changes in the oral mucosa. J Clin Exp Dent 2013;5:e187–91. https://doi.org/10.4317/jced.51121.

[30] Nagi R, Reddy-Kantharaj Y-B, Rakesh N, Janardhan-Reddy S, Sahu S. Efficacy of light based detection systems for early detection of oral cancer and oral potentially malignant disorders: Systematic review. Medicina Oral Patología Oral Y Cirugía Bucal 2016;21:e447–55. https://doi.org/10.4317/medoral.21104.

[31] Essat M, Cooper K, Bessey A, Clowes M, Chilcott JB, Hunter KD. Diagnostic accuracy of conventional oral examination for detecting oral cavity cancer and potentially malignant disorders in patients with clinically evident oral lesions: Systematic review and meta-analysis. Head Neck 2022;44:998–1013. https://doi.org/10.1002/hed.26992.

[32] García-Pola M, Pons-Fuster E, Suárez-Fernández C, Seoane-Romero J, Romero-Méndez A, López-Jornet P. Role of Artificial Intelligence in the Early Diagnosis of Oral Cancer. A Scoping Review. Cancers 2021;13:4600. https://doi.org/10.3390/cancers13184600.

[33] Warnakulasuriya S, Kerr AR. Oral Cancer Screening: Past, Present, and Future. J Dent Res 2021;100:1313–20. https://doi.org/10.1177/00220345211014795.

[34] Aubreville M, Knipfer C, Oetter N, Jaremenko C, Rodner E, Denzler J, et al. Automatic Classification of Cancerous Tissue in Laserendomicroscopy Images of the Oral Cavity using Deep Learning. Sci Rep-Uk 2017;7:11979. https://doi.org/10.1038/s41598-017-12320-8.

[35] Majumder SK, Ghosh N, Gupta PK. N2 laser excited autofluorescence spectroscopy of formalin-fixed human breast tissue. J Photochem Photobiology B Biology 2005;81:33–42. https://doi.org/10.1016/j.jphotobiol.2005.06.002.

[36] Chigurupati R, Kuriakose M. ONCOGRID: An mHealth Approach to Prevention and Early Diagnosis of Oral Cancer in Rural South India 2017.

[37] Haron N, Zain RB, Nabillah WM, Saleh A, Kallarakkal TG, Ramanathan A, et al. Mobile Phone Imaging in Low Resource Settings for Early Detection of Oral Cancer and Concordance with Clinical Oral Examination. Telemed E-Health 2017;23:192–9. https://doi.org/10.1089/tmj.2016.0128.

[38] Song B, Sunny S, Uthoff RD, Patrick S, Suresh A, Kolur T, et al. Automatic classification of dual-modalilty, smartphone-based oral dysplasia and malignancy images using deep learning. Biomed Opt Express 2018;9:5318. https://doi.org/10.1364/boe.9.005318.

[39] Franzmann EJ, Donovan MJ. Effective early detection of oral cancer using a simple and inexpensive point of care device in oral rinses. Expert Rev Mol Diagn 2018;18:837–44. https://doi.org/10.1080/14737159.2018.1523008.

[40] Martin JL. Validation of Reference Genes for Oral Cancer Detection Panels in a Prospective Blinded Cohort. Plos One 2016;11:e0158462. https://doi.org/10.1371/journal.pone.0158462.

[41] Guerrero-Preston R, Godoy-Vitorino F, Jedlicka A, Rodríguez-Hilario A, González H, Bondy J, et al. 16S rRNA amplicon sequencing identifies microbiota associated with oral cancer, human papilloma virus infection and surgical treatment. Oncotarget 2016;7:51320–34. https://doi.org/10.18632/oncotarget.9710.

[42] Zhou X, Hao Y, Peng X, Li B, Han Q, Ren B, et al. The Clinical Potential of Oral Microbiota as a Screening Tool for Oral Squamous Cell Carcinomas. Front Cell Infect Mi 2021;11:728933. https://doi.org/10.3389/fcimb.2021.728933.

[43] Kujan O, Khattab A, Oliver RJ, Roberts SA, Thakker N, Sloan P. Why oral histopathology suffers inter-observer variability on grading oral epithelial dysplasia: An attempt to understand the sources of variation. Oral Oncol 2007;43:224–31. https://doi.org/10.1016/j.oraloncology.2006.03.009.

